# Applying a computational transcriptomics-based drug repositioning pipeline to identify therapeutic candidates for endometriosis

**DOI:** 10.1101/2022.12.20.22283736

**Authors:** Tomiko T Oskotsky, Arohee Bhoja, Daniel Bunis, Brian L Le, Idit Kosti, Christine Li, Sahar Houshdaran, Sushmita Sen, Júlia Vallvé-Juanico, Wanxin Wang, Erin Arthurs, Lauren Mahoney, Lindsey Lang, Brice Gaudilliere, David K Stevenson, Juan C Irwin, Linda C Giudice, Stacy McAllister, Marina Sirota

## Abstract

Endometriosis is a common, inflammatory pain disorder comprised of disease in the pelvis and abnormal uterine lining and ovarian function that affects ∼200 million women of reproductive age worldwide and up to 50% of those with pelvic pain and/or infertility. Existing medical treatments for endometriosis-related pain are often ineffective, with individuals experiencing minimal or transient pain relief or intolerable side effects limiting long-term use - thus underscoring the pressing need for new drug treatment strategies. In this study, we applied a computational drug repurposing pipeline to endometrial gene expression data in the setting of endometriosis and controls in an unstratified manner as well as stratified by disease stage and menstrual cycle phase in order to identify potential therapeutics from existing drugs, based on expression reversal. Out of the 3,131 unique genes differentially expressed by at least one of six endometriosis signatures, only 308, or 9.8%, were in common. Similarities were more pronounced when looking at therapeutic predictions: 221 out of 299 drugs identified across the six signatures, or 73.9%, were shared, and the majority of predicted compounds were concordant across disease stage-stratified and cycle phase-stratified signatures. Our pipeline returned many known treatments as well as novel candidates. We selected the NSAID fenoprofen, the top therapeutic candidate for the unstratified signature and among the top-ranked drugs for the stratified signatures, for further investigation. Our drug target network analysis shows that fenoprofen targets PPARG and PPARA which affect the growth of endometrial tissue, as well as PTGS2 (i.e., COX2), an enzyme induced by inflammation with significantly increased gene expression demonstrated in patients with endometriosis who experience severe dysmenorrhea. NSAIDs are widely prescribed for endometriosis-related dysmenorrhea and nonmenstrual pelvic pain. Our analysis of clinical records across University of California healthcare systems revealed that while NSAIDs have been commonly prescribed to the 61,306 patients identified with diagnoses of endometriosis, dysmenorrhea, or chronic pelvic pain (36,543, 59.61%), fenoprofen was infrequently prescribed to those with these conditions (5, 0.008%). We tested the effect of fenoprofen in an established rat model of endometriosis and determined that it successfully alleviated endometriosis-associated vaginal hyperalgesia, a surrogate marker for endometriosis-related pain. These findings validate fenoprofen as a potential endometriosis therapeutic and suggest the utility of future investigation into additional drug targets identified.

## Introduction

Endometriosis is an estrogen-dependent inflammatory condition characterized by the presence of endometrial-like tissue, refluxed during menses into the pelvis or, less commonly, by hematogenous or lymphatic spread to other parts of the body. It affects over 200 million people of reproductive age worldwide and up to 50% of those with infertility^1^. The most common symptom of endometriosis is pain, and ∼ 50% of women with chronic pelvic pain have endometriosis^2^. First-line treatment for endometriosis-associated pain involves non-steroidal anti-inflammatory drugs (NSAIDs) and hormonal suppressive therapy with progestins, combined oral contraceptives, or GnRH agonists or antagonists^3^. However, these treatments are often ineffective, with nearly 19% of patients experiencing no reduction in pain and up to 59% having remaining pain^4^, thus making it essential to identify effective therapeutic candidates for endometriosis-related pain.

New drug development has been limited for endometriosis-related pain, likely due to numerous factors including the heterogeneity of the disease subtypes and presenting symptoms, and less investment globally in women’s health related disorders, including those associated specifically with menstruation and pain^5^. Also, traditional drug development is time consuming and expensive; it can take over 15 years and $1 billion to bring a new drug to market^6^. This is especially true of endometriosis due to the complexity of the disease, its etiology, and pathophysiology. It becomes essential, then, that alternate paths are pursued. Computational drug repurposing is the process of identifying novel therapeutic applications for existing compounds via computational methods. In recent years, large public datasets have been made possible by high throughput profiling technology, and efficient computation and analysis of big data have become more accessible. As a result, computational drug repurposing has gained traction as a modern innovation on traditional methodologies. Narrowing down candidates for experimental validation to existing drugs with transcriptomic profiles that suggest therapeutic effectiveness when applied to a disease mitigates the risk of failure in early stages of drug development. In addition, since every candidate is FDA approved, identified drugs have already been subjected to clinical trials and have established safety and side-effect profiles. This combination of factors vastly decreases time and cost, and shortens the path from initial development to clinical use.

One method of computational drug repurposing, pioneered by our group, uses a pattern-matching strategy to identify drugs and diseases with reversed differential gene expression profiles — where genes downregulated in a disease are upregulated by the drug treatment and vice versa. This approach relies on transcriptomics data, which can be leveraged to generate profiles of gene changes for both drugs and diseases. These profiles measure genome-wide changes in gene expression between an experimental state and a control state (in this case, a disease sample vs. a healthy control, or a drug-exposed sample vs. unperturbed cells). The hypothesis behind this method is that a drug may have a therapeutic effect on a disease if their differential gene expression profiles are opposite^7^. In the past, this method has been successfully applied to identify both known and novel treatments for inflammatory bowel disease^8^, dermatomyositis^9^, and liver cancer^10^. In addition, it has been used to identify novel therapeutics for preterm birth^11^ and COVID-19^12^, indicating the potential applications of drug repurposing to reproductive health.

In the past, transcriptomics work in endometriosis has allowed us to characterize the unique environment of endometrial lesions, which includes distinctive perivascular mural cells that promote angiogenesis and immune cell migration^13^; analyze patterns in gene expression between healthy controls and endometriosis patients, taking into account age, disease stage, menstrual cycle phase, and other clinical factors^14^; apply computational approaches to identify the individual contributions of cell subtypes to the overall endometriosis phenotype^15^; and identify specific subtypes of cells that are only enriched in control or disease tissue — proliferating uterine natural killer cells are uniquely enriched in healthy samples, and endometrial stromal cells are enriched in disease samples^16^. Transcriptomic profiling and analysis have allowed us to better understand the mechanism underlying endometriosis, and the greater availability of public datasets creates opportunities for drug repurposing.

In this study, a computational drug repurposing pipeline was applied to endometrial gene expression data in the setting of endometriosis and controls in order to identify potential therapeutics from existing drugs based on expression reversal. Moreover, we established a rat model to validate the NSAID fenoprofen, our top drug candidate, as a potential endometriosis therapeutic.

## Methods

### Study Design

The overall study overview is shown in Figure 1.

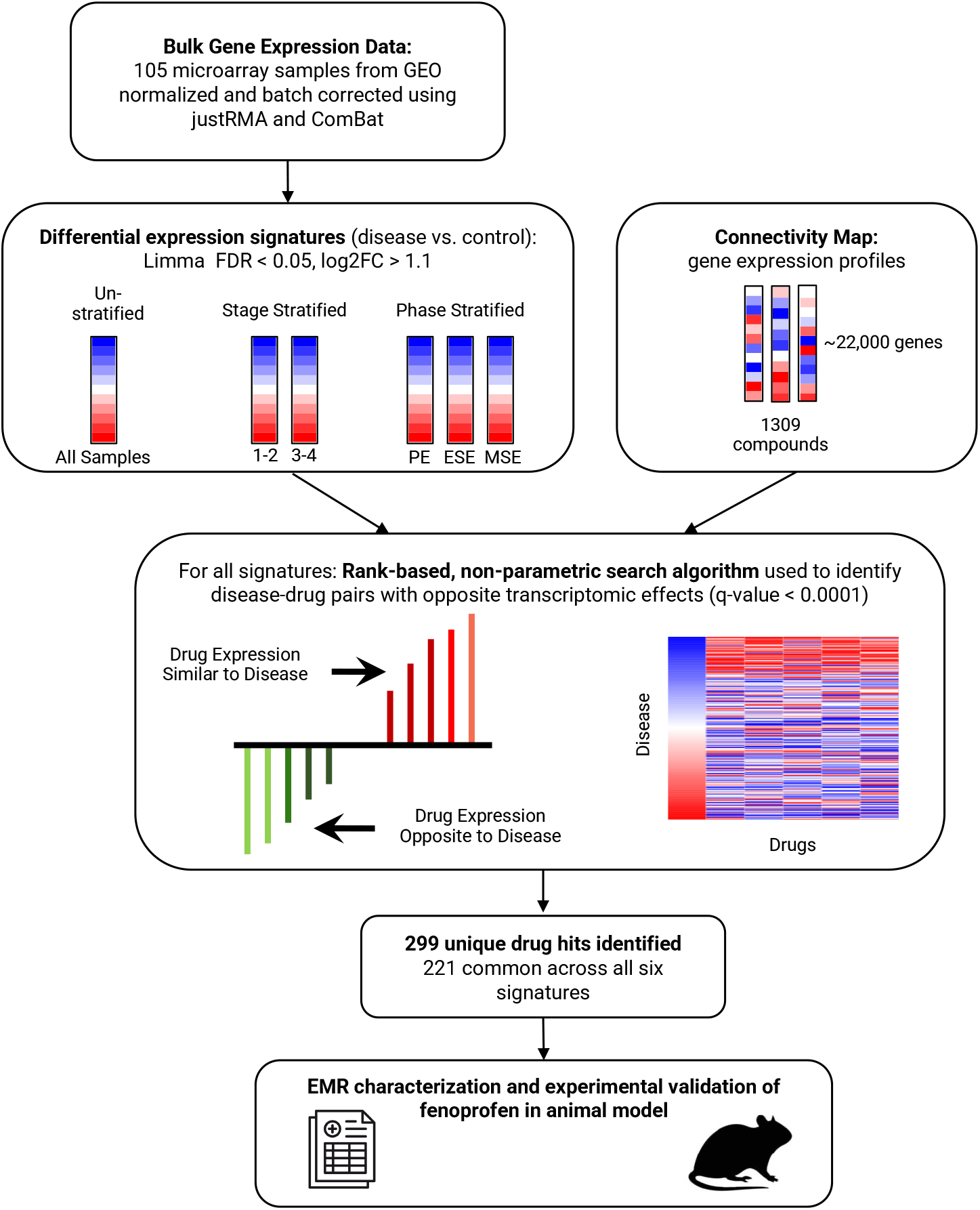

### Gene Expression Signature of Endometriosis

Microarray-based transcriptional profiling data from eutopic endometrial tissues of women, either with endometriosis or with no uterine or pelvic pathology (NUPP), were obtained from the National Center for Biotechnology Information (NCBI) Gene Expression Omnibus (GEO) Database (series accession number GSE51981) and cleaned and batch corrected as described in Bunis et al. (2021). Sample metadata were used to classify samples by lab of origin, by disease stages I-II, disease stages III-IV^17^ or NUPP control, and by cycle-phases proliferative endometrium (PE), early secretory endometrium (ESE), or mid secretory endometrium (MSE), with any samples that could not be unambiguously mapped (n=2) discarded^18^. Remaining data (n=105) were then normalized with the R package justRMA^19^ and batch corrected using the package ComBat^20^ to reduce signal associated with lab of origin, while protecting signals associated with the disease stage and the cycle phase. Using the package limma^21^), unstratified and, for sensitivity analysis, stratified differential gene expression analyses were performed on subsets of the samples: unstratified = all samples; stage-stratified = all control samples and either stage I-II or stage III-IV samples; phase-stratified = control and disease samples of the given cycle-phase (PE, ESE, or MSE). Genes passing cutoffs FDR-adjusted P-value < 0.05 and logFC > 1.1 and that were represented in the Connectivity Map (CMap) dataset from the Broad Institute^22^ were considered the significant genes for each signature.

### Computational Drug Repurposing Based on Gene Expression Profiling

To identify potential drug candidates for endometriosis, we used a nonparametric rank-based method based on differential gene expression profiles using the Kolmogorov-Smirnov statistic^10^. The hypothesis is that drugs with opposite transcriptional effects to those observed in endometriosis could potentially have a therapeutic effect in treating endometriosis. On the drug side, CMap was used to obtain gene expression profiles from various cell lines treated with small-molecule drugs. The CMap dataset has gene expression profiles (∼22,000 genes) for 1,309 small-molecule drug compounds cultured in up to 5 different cell lines^22^. For endometriosis, differential gene expression signatures were generated as described using six different stratifications: unstratified (all samples), stratified by stage (stages I-II or stages III-IV), and stratified by phase (PE, ESE, or MSE).

For each endometriosis disease signature, reversal scores were calculated for each drug in the CMap dataset based on the relationship between the gene expression profiles for the disease relative to the drug. A negative score indicated reverse profiles between the drug and disease signature, where upregulated genes in the disease signature were downregulated in the drug signature and vice versa. A positive score indicated the opposite — similar profiles between the drug and disease signature. For drugs with multiple gene expression profiles from different cell lines or concentrations, we kept the profile with the largest reversal effect, or most negative score. Permutation analysis was carried out to assess significance, and drug hits with q-values < 0.0001 or reversal scores < 0 (indicating signature reversal) were examined further.

### Electronic Health Record Analysis

The study was approved by the University of California, San Francisco, institutional review board and considered secondary research for which consent is not required. Patients with endometriosis, chronic pelvic pain, or dysmenorrhea who were prescribed (a) any NSAID and (b) fenoprofen were identified from the UC Data Discovery Portal’s UC-wide OMOP-based EMR database, which includes clinical data from over 8 million patients from January 1, 2012 to July 30, 2022 at UC San Francisco, UC Davis, UC Irvine, UC Los Angeles, and UC San Diego. During August 2022, patients from these five UC institutions with endometriosis, chronic pelvic pain, or dysmenorrhea were identified by inclusion criteria of having a self- or provider-identified sex of female with at least one OMOP concept id for endometriosis (OMOP concept ids 4211992, 37117191, 4072148, 4146995, 4264439, 4182703, 36713393, 4288543, 4307585, 4176409, 4051345, 4058381, 4200841, 4260818, 194420, 4272614, 4132140, 37209400, 197033, 4222798, 4317964, 4127413, 4019817, 139882, 37209399, 199881, 4189364, 36713394, 4276944, 37119080, 194421, 4230333, 46273242, 42536674, 36717630, 433527, 37110261, 37110262, 4224161, 4195507, 37209188, 4034016, 44806162, 37396113, 44806981, 4167725, 42737048, 2109446, 42737049, 2109445, 2109444, 4306918, 4202522, and 4270918), for chronic pelvic pain (OMOP concept ids 4133035, 4034006, and 42534971), or for dysmenorrhea (OMOP concept ids 4137754, 4159586, 4117874, and 194696). Among these patients, we identified individuals who were ever (a) prescribed at least one of the following NSAIDs: ibuprofen, naproxen, celecoxib, diclofenac, etodolac, indomethacin, piroxicam, sulindac, oxaprozin, meloxicam, diflunisal, ketorolac, meclofenamate, nabumetone, salsalate, and fenoprofen, and (b) prescribed fenoprofen.

### Animal Model Validation

#### Subjects and vaginal cytology

Animal subjects were 24 virgin female Sprague Dawley rats (175-200 g at arrival; Charles River, Raleigh, NC). All rats were single housed in plastic cages lined with chip bedding and *ad libitum* access to rodent chow and water. Housing conditions were environmentally controlled (room temperature ∼22°C, 12-hour light/dark cycle, lights on at 07:00). Reproductive status was determined by vaginal lavage performed ∼2 hours after lights on using traditional nomenclature for the 4 estrous stages of proestrus, estrus, metestrus, and diestrus^23^. The study and procedures were approved by the Emory University Institutional Animal Care and Use Committee (IACUC) as protocol #2021000201. All laboratory animal experimentation adhered to the NIH Guide for the Care and Use of Laboratory Animals.

#### Endometriosis model (ENDO)

The ENDO model was performed as previously described^24^. At 18-20 weeks of age, in diestrus, rats were anesthetized intraperitoneally with a mixture of ketamine hydrochloride (73 mg/kg) and xylazine (8.8 mg/kg) and placed on a heating pad to maintain body temperature (∼37°C). An off-midline (left side) incision was made through the skin and muscle layer to expose the pelvic and abdominal organs. An □1-cm segment of mid-left uterine horn was excised and placed in warm saline. Four, 2 × 2-mm pieces of excised uterus were sewn onto alternate mesenteric arteries that supply the caudal small intestine starting from the caecum using 4.0 nylon sutures. After it was confirmed that there was no bleeding in the abdominal cavity, the muscle layer and skin incision were closed with chromic gut and non-absorbable suture, respectively. Rats were monitored closely during recovery; the postoperative recovery period was uneventful.

#### Behavioral assessment of vaginal nociception

The behavioral training and testing procedures were performed as previously described^25^. Rats were trained to perform an escape response to terminate vaginal distention produced by an inflatable latex balloon. During each testing session, eight different distention volumes were delivered three times each in random order at intervals of ∼60 seconds, and percent escape response to each volume was assessed.

##### Behavioral apparatus and stimulator

The training and testing apparatus was a grill-floored Plexiglas® chamber allowing movement but preventing the rat from turning around. In the front of the chamber, a hollow tube is extended containing a light-emitting diode and photo sensor. When a rat extends her nose into this tube, the light beam is broken, and the stimulus is terminated constituting an “escape response”. An opening in the rear of the chamber allows the catheter (attached to the vaginal stimulator) to connect to a computer-controlled stimulus-delivery device.

The vaginal stimulator is a small latex balloon (∼ 10mm × 1.5 mm uninflated) tied to a catheter with silk suture. Prior to testing, the uninflated balloon is lubricated with K-Y® jelly and inserted into the mid-vaginal canal. The vaginal canal is then distended by delivery of different volumes of water to the balloon.

##### Behavioral training

Rats were first allowed to acclimate to the testing chamber 10 minutes daily for 3-4 days. Then, over a time period of 4-6 weeks, rats were trained to: face forward in the testing chamber without turning around (box training: 3-4 sessions, consecutive days), perform an “escape response” by extending their head into a hollow tube to interrupt a light beam (tail pinch training: 4-8 sessions, non-consecutive days), and perform an identical “escape response” to terminate vaginal distention stimuli (balloon training: 3-5 sessions, non-consecutive days).

##### Behavioral testing

Once trained, 1-hour long testing sessions consisted of 24 computer-controlled escape trials run at ∼1-minute intervals (range 50-70 seconds). Each trial consisted of rapid inflation of the balloon (1 mL/s) to a fixed volume until the rat made an “escape response” or 15 seconds had elapsed, when the balloon rapidly deflated (0.5 mL/s). Eight different distention volumes (0.15, 0.30, 0.40, 0.55, 0.70, 0.80, 0.90 mL), including a control volume (0.01mL), were delivered to the balloon three times each in random order. The computer recorded the stimulus and escape response for each trial. The maximum latency of 15 seconds was considered no response. Testing sessions were run 3 times/week, on non-consecutive days.

#### Experimental groups

There were four groups of rats analyzed: Group 1: ENDO, fenoprofen (30 mg/kg/day, p.o); Group 2: ENDO, ibuprofen (30 mg/kg/day, p.o.) (positive control); Group 3: ENDO, no treatment (negative control); and Group 4: no ENDO, no treatment (negative control). Ibuprofen was selected as our positive control as it is a commonly used analgesic agent for rodents and has been effectively used in pain studies in rodents (e.g. inflammatory pain models)^26–28^. In all groups, vaginal nociception was behaviorally assessed over three chronological testing periods as follows: (i) testing period 1: an initial baseline period of 8 weeks, (ii) testing period 2: a post-ENDO or middle-testing period of 10 weeks, and (iii) testing period 3: a post-treatment or late-testing period of 4 weeks. Data for rats in control Groups 3 and 4 (n=3/group) were retrieved and reanalyzed from an earlier study^29^.

## Results

### Computational identification of drug repurposing candidates

The gene expression data were derived from samples collected from women with minimal to mild (stage I–II) or more advanced (stage III-IV) endometriosis, and those without uterine or pelvic pathology (control) in proliferative, early secretory, and mid-secretory phases of the menstrual cycle. General cohort characteristics are shown in Supplementary Table 1 and provided previously in greater detail^18^. The numbers of significant differentially expressed genes from unstratified, stage-stratified (i.e., stage I–II or stage III-IV), and phase-stratified (i.e., PE, MSE, or ESE) comparisons of patients with endometriosis to patients in the control cohort are represented in Table 1, with the specific differentially expressed genes represented in Supplementary Tables 2A-F. Out of the 3,131 unique genes differentially expressed by at least one endometriosis signature and represented in the CMap database, only 308, or 9.8%, were common across all six signatures (Figure 2).

**Table 1:**
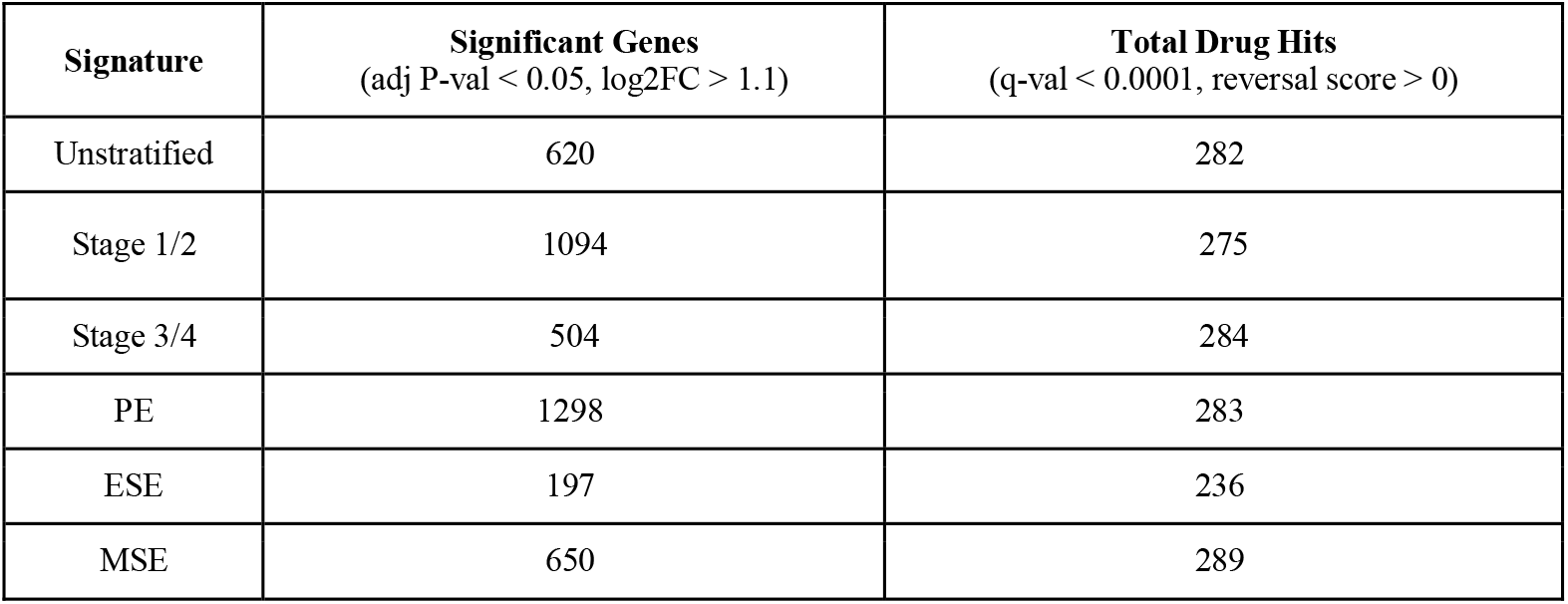
Significant genes that were also represented in CMAP and total drug hits

| **Signature** | **Significant Genes**<br>(adj P-val < 0.05, log2FC > 1.1) | **Total Drug Hits**<br>(q-val < 0.0001, reversal score > 0) |
| --- | --- | --- |
| Unstratified | 620 | 282 |
| Stage 1/2 | 1094 | 275 |
| Stage 3/4 | 504 | 284 |
| PE | 1298 | 283 |
| ESE | 197 | 236 |
| MSE | 650 | 289 |

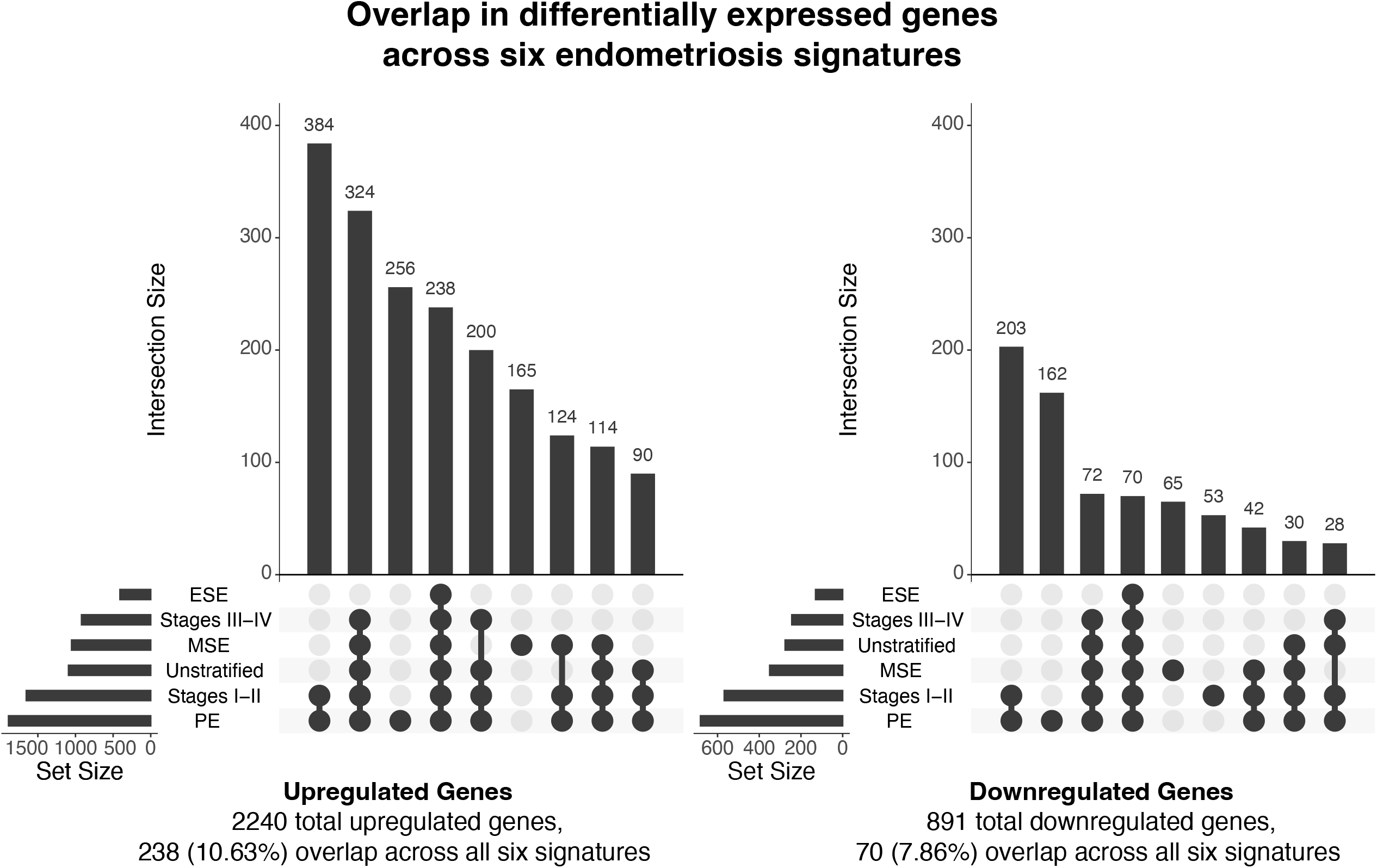

By analyzing via our drug repositioning pipeline, the unstratified and stratified differential gene expression signatures with the drug signatures from the CMap dataset, 236-289 drug candidates were determined per signature and 299 unique drugs that significantly reversed the expression profiles of the disease. Similarities across the six signatures were more pronounced for drug candidates than for differentially expressed genes: 221 out of 299 drugs, or 73.9%, were common to every signature (Figure 3A). Of the 221 drugs common across all six signatures, many returned high reversal scores across the board, suggesting that these compounds could be used to treat different stages of endometriosis and across every cycle phase. As there was consistency in the majority of the drug candidates from the unstratified and stratified signatures, we moved forward based solely on the unstratified endometriosis signature.

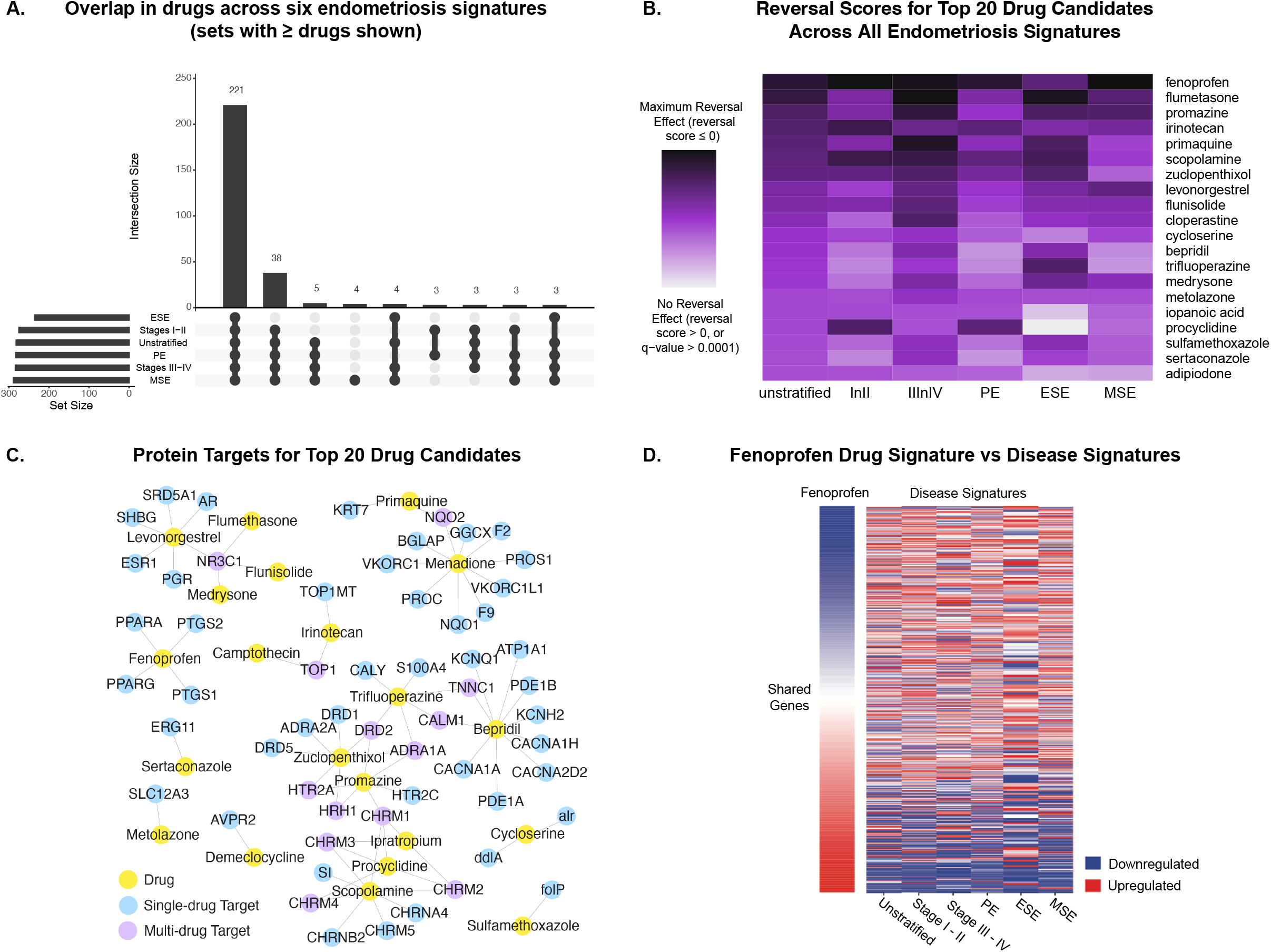

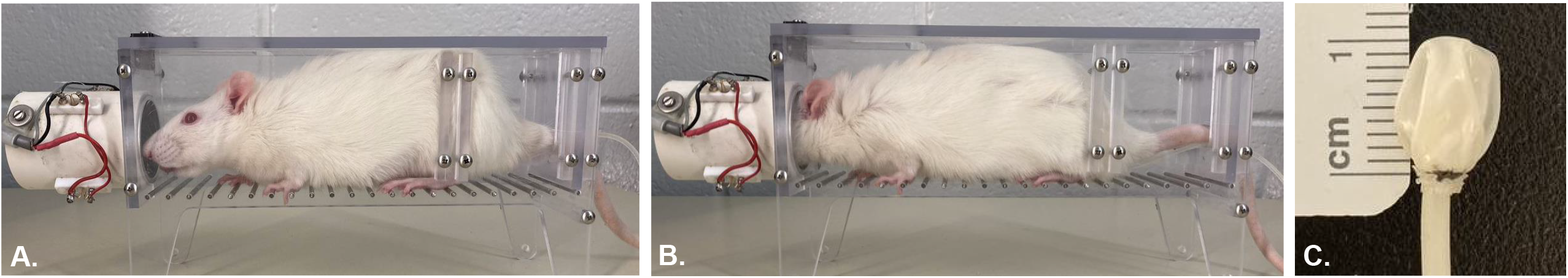

Several of the drug candidates identified are from classes of medications used to treat endometriosis. Levonorgestrel, among the top 10 drug candidates, is a progestin recommended for treating endometriosis^30^. Also among the identified drug candidates are non-steroidal anti-inflammatory drugs (NSAIDs) such as acetylsalicylic acid (commonly known as aspirin), mefenamic acid, indomethacin, naproxen, and diclofenac, which are frequently recommended to alleviate pain and inflammation in dysmenorrhea patients. The NSAID ibuprofen, its isomer dexibuprofen, and the COX-2 selective inhibitor NSAIDs celecoxib, rofecoxib, and valdecoxib are not among our predicted therapeutic candidates since ibuprofen is not represented in CMAP, and dexibuprofen as well as the COX-2 inhibitors are represented in CMAP but are filtered out during pre-processing due to profile inconsistencies^10^. A heatmap of the top 20 drug candidates and their reversal scores for the six endometriosis signatures is shown in Figure 3B and demonstrates consistency of predictions across the signatures.

Using the DrugBank database^31^, we were able to identify several proteins targeted by our top 20 drug candidates, including 13 that were targeted by two or more of identified drugs. These interactions were used to construct a network, which can be used to visualize the unique and shared interactions between the top 20 drug candidates and their protein targets (Figure 3C). Out of the proteins displayed in the network, several were found to have a link to endometriosis. Peroxisome proliferator activated receptors gamma and alpha (PPARG and PPARA), which are commonly targeted by NSAID drugs including fenoprofen, can impede the growth of endometrial tissue when activated^32,33^. Prostaglandin-endoperoxidase synthase 2 (PTGS2) gene expression has been found to be significantly increased in ectopic and eutopic endometrium of women with endometriosis compared to women without this condition^34,35^. Moreover, among endometriosis patients, PTGS2 expression in eutopic endometrium has been shown to be significantly greater in women with higher pain scores for dysmenorrhea^36^. Dopamine receptor type-2 (DRD2) polymorphisms have been identified in patients with endometriosis, and treatment with DRD2 agonists has been associated with the disappearance or decrease in size of peritoneal endometriotic lesions^37,38^. Increased gene expression of steroid 5 alpha-reductase 1 (SRD5A1) has been found in ovarian endometriosis compared to normal endometrium^39^. In addition, the nuclear receptor proteins NR3C1 (nuclear receptor subfamily 3 group C member 1), AR (androgen receptor), PR (progesterone receptor), and ESR1 (estrogen receptor 1) are expressed in endometrial cells^40,41^.

Through leveraging this approach, fenoprofen, an NSAID commonly used to treat pain and arthritis, was identified as the top drug candidate for the unstratified signature and among the top seven drug candidates for the stratified signatures. When visualizing the gene regulation of the six input endometriosis signatures and fenoprofen, the overall reversal pattern can be observed (Figure 3D). As fenoprofen had the highest reversal score of our drug candidates and belongs to a gold standard treatment category of drugs for endometriosis, our validation efforts herein were focused on this medication.

### Electronic Health Record Analysis

From the analysis of electronic medical records (EMR) across five University of California (UC) healthcare institutions (UC San Francisco (UCSF), UC Davis, UC Irvine, UC Los Angeles, and UC San Diego), there were a total of 61,306 patients with endometriosis, chronic pelvic pain, or dysmenorrhea, among whom 36,543 (59.61%) had a prescription for an NSAID and 5 (0.008%) had a prescription for fenoprofen (Table 2). For the individual UC healthcare institutions, among the 12,476 patients at UCSF with endometriosis, chronic pelvic pain, or dysmenorrhea, 7,752 (62.14%) had a prescription for an NSAID and 1 (0.008%) had a prescription for fenoprofen; among the 10,040 patients at UC Davis with endometriosis, chronic pelvic pain, or dysmenorrhea, 6163 (61.38%) had a prescription for an NSAID and 0 (0.000%) had a prescription for fenoprofen; among the 5,694 patients at UC Irvine with endometriosis, chronic pelvic pain, or dysmenorrhea, 3,545 (62.26%) had a prescription for an NSAID and 0 (0.000%) had a prescription for fenoprofen; among the 23017 patients at UC Los Angeles with endometriosis, chronic pelvic pain, or dysmenorrhea, 13,041 (56.66%) had a prescription for an NSAID and 3 (0.013%) had a prescription for fenoprofen; and among the 10,079 patients at UC San Diego with endometriosis, chronic pelvic pain, or dysmenorrhea, 6,042 (59.95%) had a prescription for an NSAID and 1 (0.010%) had a prescription for fenoprofen (Table 2).

**Table 2.** Prevalence of NSAID prescriptions: Patients with endometriosis, chronic pelvic pain, or dysmenorrhea and prescribed (a) any NSAID and (b) fenoprofen at five UC Health Care Institutions (UCSF, UCD, UCI, UCLA, UCSD)

| **Institution** | **Patients with endometriosis, chronic pelvic pain, or dysmenorrhea** | **Patients with endometriosis, chronic pelvic pain, or dysmenorrhea and prescribed any NSAID (%)** | **Patients with endometriosis, chronic pelvic pain, or dysmenorrhea and prescribed fenoprofen (%)** |
| --- | --- | --- | --- |
| UCSF | 12476 | 7752 (62.14%) | 1 (0.008%) |
| UC Davis | 10040 | 6163 (61.38%) | 0 (0%) |
| UC Irvine | 5694 | 3545 (62.26%) | 0 (0%) |
| UC Los Angeles | 23017 | 13041 (56.66%) | 3 (0.013%) |
| UC San Diego | 10079 | 6042 (59.95%) | 1 (0.010%) |
| Total | 61306 | 36543 (59.61%) | 5 (0.008%) |

### Animal Model Validation

To validate the in-silico findings, we used an established animal model of endometriosis that produces vaginal hyperalgesia, a surrogate marker for endometriosis-related pain. In this model, uterine pieces are autotransplanted onto mesenteric abdominal arteries which form vascularized cyst-like structures. Vaginal hyperalgesia, or an increase in vaginal nociception, develops and stabilizes by ten weeks in this model^29^. Vaginal nociception was assessed as an escape response to a noxious stimulus, a water filled balloon. Escape response was measured as a function of vaginal balloon distention volume. Fenoprofen was dosed orally for four weeks. As controls, ibuprofen was orally dosed or no treatment was delivered. As an additional control, rats with no endometriosis received no treatment. In all four groups, vaginal nociception was assessed and compared over three testing periods (i) an initial baseline period of eight weeks (ii) a post-endo or middle-testing period of ten weeks, and (iii) a post-treatment or late-testing period of four weeks.

Responses among fenoprofen (30 mg/kg/day, p.o.) were significantly increased during the post-endo surgery period compared to the baseline period, when volumes of 0.15, 0.3, 0.4, 0.55, 0.7, and 0.8 mL of water were delivered (Mann Whitney U test, Bonferroni-corrected p-value threshold of 0.05. Figure 5A, Table 3). During the post-treatment period, escape responses were significantly decreased compared to the post-endo surgery period when volumes of 0.15, 0.3, 0.4, 0.55, 0.7, and 0.8 mL of water were delivered (Mann Whitney U test, Bonferroni-corrected p-value threshold of 0.05. Figure 5A, Table 3). No statistically significant difference was found in the escape responses between the baseline period and the post-treatment period for any volume of water delivered to the fenoprofen treated subjects (Mann Whitney U test, Bonferroni-corrected p-value threshold of 0.05. Figure 5A, Table 3).

**Table 3.** Responses with Fenoprofen treatment. Median escape response with interquartile range (IQR) for each delivered volume (0.01, 0.15,0.30, 0.40, 0.55, 0.70, 0.80, and 0.90 mL) during the baseline, post-endo surgery, and post-treatment periods, with Bonferroni-corrected p-values from Mann-Whitney *U* test for baseline period vs post-endo surgery period, post-endo surgery vs post-treatment period, and baseline period vs post-treatment period. * denotes Bonferroni-corrected p-values below significance threshold of 0.05.

| Volume | Baseline period (BL) median | BL IQR | Post-Endo period (PE) median | PE IQR | Post-Treatment period (PT) median | PT IQR | Baseline period vs Post-Endo period MWU test Bonferroni-adjusted p-values | Post-Endo period vs Post-Treatment period MWU test Bonferroni-adjusted p-values | Baseline period vs Post-Treatment period MWU test Bonferroni-adjusted p-values |
| --- | --- | --- | --- | --- | --- | --- | --- | --- | --- |
| 0.01 | 0 | (0-0) | 0 | (0-0) | 0 | (0-0) | 1 | 1 | 1 |
| 0.15 | 0 | (0-0) | 33.3 | (0-66.6) | 0 | (0-0) | 2.7E-04 * | 2.3E-03 * | 0.61 |
| 0.3 | 0 | (0-25) | 33.3 | (33.3-66.6) | 0 | (0-33.3) | 2.0E-05 * | 3.0E-06 * | 1 |
| 0.4 | 0 | (0-33.3) | 66.6 | (33.3-91.7) | 16.65 | (0-33.3) | 4.0E-06 * | 1.8E-05 * | 1 |
| 0.55 | 33.3 | (8.3-33.3) | 83.3 | (66.6-100) | 33.3 | (33.3-66.6) | 2.7E-07 * | 7.9E-07 * | 0.16 |
| 0.7 | 66.6 | (33.3-66.6) | 100 | (100-100) | 66.6 | (66.6-100) | 1.5E-03 * | 4.4E-02 * | 0.97 |
| 0.8 | 66.6 | (66.6-100) | 100 | (100-100) | 100 | (66.6-100) | 4.8E-05 * | 5.8E-03 * | 1 |
| 0.9 | 100 | (100-100) | 100 | (100-100) | 100 | (100-100) | 0.34 | 0.80 | 1 |

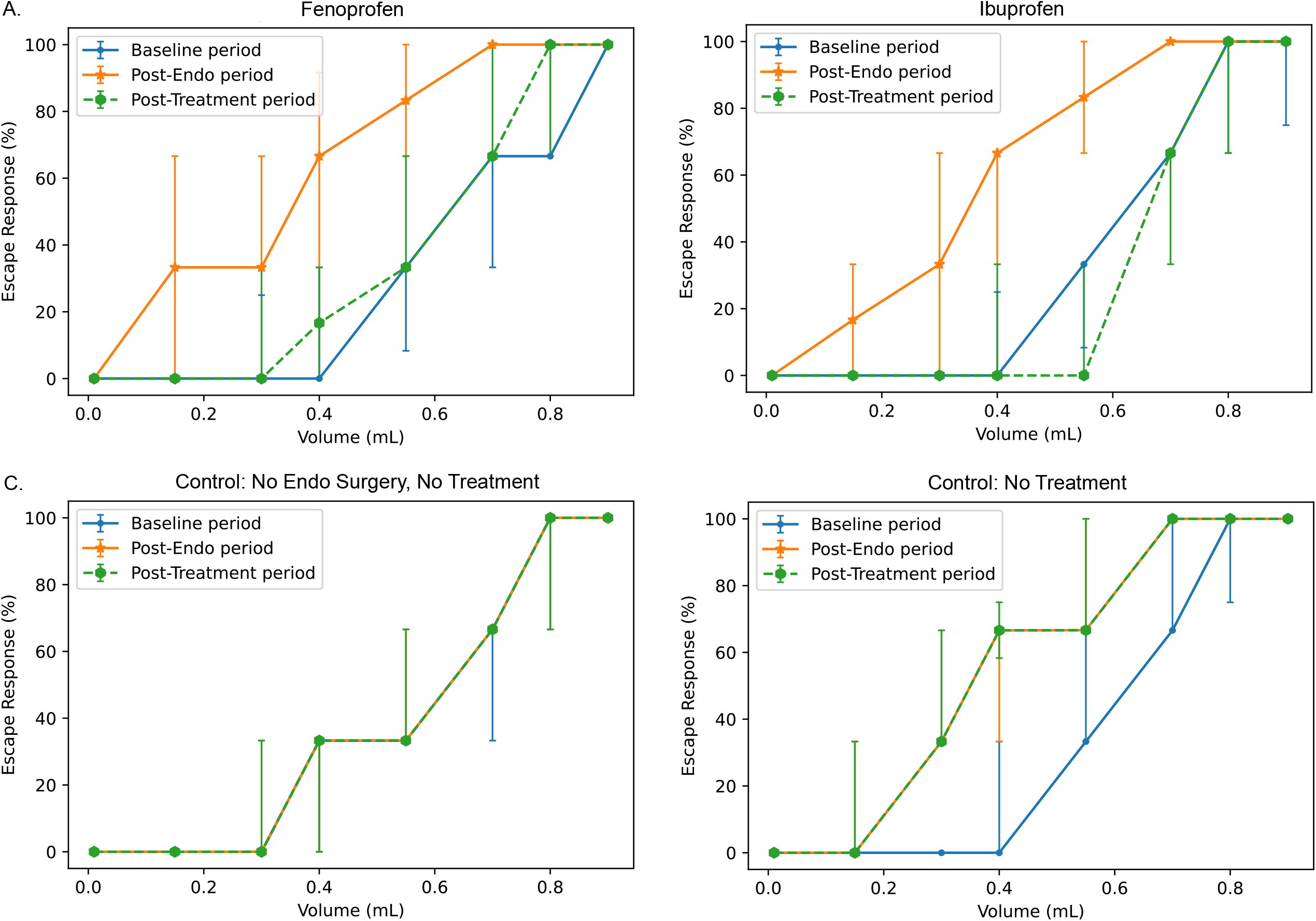

Similarly, among ibuprofen (30 mg/kg/day, p.o.) treated animals, escape responses were significantly increased during the post-endo surgery period compared to the baseline period, when volumes of 0.15, 0.3, 0.4, 0.55, 0.7, and 0.8 mL of water were delivered (Mann Whitney U test, Bonferroni-corrected p-value threshold of 0.05. Figure 5B, Table 4). During the post-treatment period, escape responses were significantly decreased compared to the post-endo surgery period, when volumes of 0.15, 0.3, 0.4, 0.55, 0.7, and 0.8 mL of water were delivered (Mann Whitney U test, Bonferroni-corrected p-value threshold of 0.05. Figure 5B, Table 4). No statistically significant difference was found in the escape responses between the baseline period and the post-treatment period for any volume of water delivered to the Ibuprofen treated subjects (Mann Whitney U test, Bonferroni-corrected p-value threshold of 0.05. Figure 5B, Table 4).

**Table 4.** Responses with Ibuprofen treatment (positive control). Median escape response with interquartile range (IQR) for each delivered volume (0.01, 0.15,0.30, 0.40, 0.55, 0.70, 0.80, and 0.90 mL) during the baseline, post-endo surgery, and post-treatment periods, with Bonferroni-corrected p-values from Mann-Whitney *U* test for baseline period vs post-endo surgery period, post-endo surgery period vs post-treatment period, and baseline period vs post-treatment period. * denotes Bonferroni-corrected p-values below significance threshold of 0.05.

| Volume | Baseline period (BL) median | BL IQR | Post-Endo period (PE) median | PE IQR | Post-Treatment period (PT) median | PT IQR | Baseline period vs Post-Endo period MWU Bonferroni-adjusted p-values | Post-Endo period vs Post-Treatment period MWU test Bonferroni-adjusted p-values | Baseline period vs Post-Treatment period MWU test Bonferroni-adjusted p-values |
| --- | --- | --- | --- | --- | --- | --- | --- | --- | --- |
| 0.01 | 0 | (0-0) | 0 | (0-0) | 0 | (0-0) | 1 | 1 | 1 |
| 0.15 | 0 | (0-0) | 16.65 | (0-33.3) | 0 | (0-0) | 3.5E-03 * | 2.3E-03 * | 1 |
| 0.3 | 0 | (0-0) | 33.3 | (0-66.6) | 0 | (0-0) | 2.0E-03 * | 9.7E-04 * | 1 |
| 0.4 | 0 | (0-25) | 66.6 | (33.3-66.6) | 0 | (0-33.3) | 1.5E-05 * | 2.9E-06 * | 1 |
| 0.55 | 33.3 | (8.3-33.3) | 83.3 | (66.6-100) | 0 | (0-33.3) | 2.5E-07 * | 3.4E-08 * | 1 |
| 0.7 | 66.6 | (66.6-66.6) | 100 | (100-100) | 66.6 | (33.3-66.6) | 5.8E-05 * | 5.1E-06 * | 1 |
| 0.8 | 100 | (66.6-100) | 100 | (100-100) | 100 | (66.6-100) | 3.9E-03 * | 1.8E-03 * | 1 |
| 0.9 | 100 | (74.9-100) | 100 | (100-100) | 100 | (100-100) | 0.12 | 0.80 | 1 |

Among animals that received neither endo surgery nor treatment, no statistically significant difference was found in the escape responses between the baseline and post-endo surgery periods, the post-endo surgery and post-treatment periods, or the baseline and post-treatment periods when any volume of water was delivered (Mann Whitney U test, Bonferroni-corrected p-value threshold of 0.05. Figure 5C, Table 5).

**Table 5.** Responses with no endo surgery and no treatment (negative control). Median escape response with interquartile range (IQR) for each delivered volume (0.01, 0.15,0.30, 0.40, 0.55, 0.70, 0.80, and 0.90 mL) during the baseline, post-endo surgery, and post-treatment periods, with Bonferroni-corrected p-values from Mann-Whitney *U* test for baseline period vs post-endo surgery period, post-endo surgery vs post-treatment period, and baseline period vs post-treatment period. * denotes Bonferroni-corrected p-values below significance threshold of 0.05.

| Volume | Baseline period (BL) median | BL IQR | Post-Endo period (PE) median | PE IQR | Post-Treatment period (PT) median | PT IQR | Baseline period vs Post-Endo period MWU test Bonferroni-adjusted p-values | Post-Endo period vs Post-Treatment period MWU test Bonferroni-adjusted p-values | Baseline period vs Post-Treatment period MWU test Bonferroni-adjusted p-values |
| --- | --- | --- | --- | --- | --- | --- | --- | --- | --- |
| 0.01 | 0 | (0-0) | 0 | (0-0) | 0 | (0-0) | 1 | 1 | 1 |
| 0.15 | 0 | (0-0) | 0 | (0-0) | 0 | (0-0) | 1 | 1 | 1 |
| 0.3 | 0 | (0-0) | 0 | (0-33.3) | 0 | (0-33.3) | 1 | 1 | 1 |
| 0.4 | 33.3 | (0-33.3) | 33.3 | (0-33.3) | 33.3 | (0-33.3) | 1 | 1 | 1 |
| 0.55 | 33.3 | (33.3-33.3) | 33.3 | (33.3-66.6) | 33.3 | (33.3-66.6) | 1 | 1 | 1 |
| 0.7 | 66.6 | (33.3-66.6) | 66.6 | (66.6-66.6) | 66.6 | (66.6-66.6) | 1 | 1 | 1 |
| 0.8 | 100 | (66.6-100) | 100 | (66.6-100) | 100 | (66.6-100) | 1 | 1 | 1 |
| 0.9 | 100 | (100-100) | 100 | (100-100) | 100 | (100-100) | 1 | 1 | 1 |

Among animals that received endo surgery but no treatment, escape responses were significantly increased during the post-endo surgery period compared to the baseline period, when volumes of 0.15, 0.3, 0.4, 0.55, and 0.7 mL of water were delivered (Mann Whitney U test, Bonferroni-corrected p-value threshold of 0.05. Figure 5D, Table 6). During the post-treatment period, escape responses were also significantly increased compared to the baseline period when volumes of 0.3, 0.4, and 0.55 mL of water were delivered (Mann Whitney U test, Bonferroni-corrected p-value threshold of 0.05. Figure 5D, Table 6). No statistically significant difference was found in the escape responses between the post-endo surgery period and the post-treatment period for any volume of water delivered to these subjects (Mann Whitney U test, Bonferroni-corrected p-value threshold of 0.05. Figure 5D, Table 6).

**Table 6.** Responses with no treatment (positive control). Median escape response with interquartile range (IQR) for each delivered volume (0.01, 0.15,0.30, 0.40, 0.55, 0.70, 0.80, and 0.90 mL) during the baseline, post-endo surgery, and post-treatment periods, with Bonferroni-corrected p-values from Mann-Whitney *U* test for baseline period vs post-endo surgery period, post-endo surgery period vs post-treatment period, and baseline period vs post-treatment period. * denotes Bonferroni-corrected p-values below significance threshold of 0.05.

| Volume | Baseline period (BL) median | BL IQR | Post-Endo period (PE) median | PE IQR | Post-Treatment period (PT) median | PT IQR | Baseline period vs Post-Endo period MWU period Bonferroni-adjusted p-values | Post-Endo period vs Post-Treatment period MWU period Bonferroni-adjusted p-values | Baseline period vs Post-Treatment period MWU period Bonferroni-adjusted p-values |
| --- | --- | --- | --- | --- | --- | --- | --- | --- | --- |
| 0.01 | 0 | (0-0) | 0 | (0-0) | 0 | (0-0) | 1 | 1 | 1 |
| 0.15 | 0 | (0-0) | 0 | (0-33.3) | 0 | (0-33.3) | 0.02 * | 1 | 0.06 |
| 0.3 | 0 | (0-0) | 33.3 | (33.3-66.6) | 33.33 | (33.3-66.6) | 4E-05 * | 1 | 2E-06 * |
| 0.4 | 0 | (0-33.3) | 66.6 | (33.3-66.7) | 66.6 | (58.3-75) | 4E-06 * | 1 | 2E-05 * |
| 0.55 | 33.3 | (33.3-66.6) | 66.66 | (66.6-100) | 66.63 | (66.6-100) | 6E-06 * | 1 | 2E-05 * |
| 0.7 | 66.6 | (66.6-100) | 100 | (100-100) | 100 | (100-100) | 7E-03 * | 1 | 0.07 |
| 0.8 | 100 | (75-100) | 100 | (100-100) | 100 | (100-100) | 0.43 | 1 | 0.60 |
| 0.9 | 100 | (100-100) | 100 | (100-100) | 100 | (100-100) | 1 | 1 | 1 |

## Discussion

Endometriosis is an estrogen-dependent inflammatory disorder, with both local (pelvic) and systemic components that commonly contribute to pelvic pain and infertility^42–44^. Therapies for pain include surgical resection of disease and/or medical approaches mostly aimed at reducing ovarian estrogen action or production. Unfortunately, ∼50% of patients need repeat surgery within 5 years or recurrent symptoms, and medical therapies are either ineffective or promote intolerable side effects that limit their long-term use^1,4^. Recent FDA approval of new drugs for endometriosis (e.g., GnRH antagonists Elagolix and Myfembree) brings hope for those who suffer from this disease^45,46^; however, off-target effects (e.g., bone density) await long-term, post-marketing studies. Thus, there is a pressing need for novel drug discovery to improve patient symptoms and quality of life. Our study identified several existing drugs with potential therapeutic applications to endometriosis using a transcriptomics-based computational drug repurposing approach. The differential gene expression profiles for endometriosis that were unstratified as well as, for sensitivity analysis, stratified by disease stage and menstrual cycle phase were compared with the profiles of several small molecule compounds tested on human cell lines, yielding 299 unique drug hits with significant (q-value < 0.0001) reversal effects. We found that therapeutic predictions were relatively consistent across stage and cycle phase with 221 shared predictions.

When categorized by drug class/ATC code, two prominent categories for the predicted therapeutics were anti-inflammatory drugs and sex hormones; drugs from both categories have extensively been used to treat endometriosis^3^. Several drugs that the pipeline returned are current gold standard treatments, such as levonorgestrel, mefenamic acid, acetylsalicylic acid (aspirin), and naproxen; others were novel candidates.

From the drugs identified by the computational drug repurposing approach, we chose fenoprofen for further validation since it returned the highest reversal score and belongs to a class of drugs (NSAIDs) that is a current first-line treatment for endometriosis. Fenoprofen is a medication available by prescription only and has been in clinical use since this drug was approved by the FDA in 1976^47^ and is indicated for the relief of mild to moderate pain in adults and, in particular, relief of signs and symptoms of rheumatoid arthritis and osteoarthritis^48^. In our analysis of the electronic medical records across five University of California healthcare institutions, we found that while NSAIDs have been commonly prescribed (56.66% to 62.26%) for patients with endometriosis, chronic pelvic pain, or dysmenorrhea diagnosis, the NSAID fenoprofen was prescribed for the minority (0% to 0.013%) of patients with these conditions.

We tested the NSAID fenoprofen in a rat model of endometriosis that displays vaginal hyperalgesia, a surrogate marker for endometriosis-related pain. We determined that oral treatment with fenoprofen significantly alleviated endometriosis-associated vaginal hyperalgesia, similar to oral ibuprofen treatment. In endometriosis rats with no treatment, vaginal hyperalgesia was maintained, which confirmed that the alleviation of hyperalgesia observed in the treatment groups was not due to additional vaginal nociceptive testing post-endometriosis. In rats with no endometriosis and no treatment, no significant changes in vaginal nociception occurred, which suggests that any observed changes in the other groups were not due to lengthy vaginal nociceptive testing alone. Overall, these findings support fenoprofen as a potential therapeutic for endometriosis-associated pain.

NSAIDs prevent or reduce production of prostaglandins, which in turn can help relieve pain from endometriosis. While NSAIDs are a commonly prescribed, first-line treatment of endometriosis, there is currently limited evidence to support the effectiveness of any NSAID over another for endometriosis pain relief^49^. Our findings suggest that the NSAID fenoprofen, currently infrequently prescribed for endometriosis, may be an effective treatment for individuals with this condition, although this would need further validation in patient cohorts. Our drug target network analysis for the top drug candidates shows that PPARG and PPARA, which impede the growth of endometrial tissue when activated, are both targeted by fenoprofen^32,33^. In addition, our network analysis showed that fenoprofen targets the enzymes PTGS1 and PTGS2 (i.e., COX1 and COX2, respectively). PTGS1 and PTGS2 have been shown to be inhibited to varying degrees in blood and gastric tissue by the available NSAIDs^50^, but any differences in the degree of inhibition of PTGS1 and PTGS2 in endometrial tissue by fenoprofen and other NSAIDs have not yet been demonstrated. PTGS1 is a constitutively expressed enzyme and involved in maintaining cell homeostasis^51^. In contrast, PTGS2 is an enzyme uncommonly expressed normally, but is induced during inflammation as well as cell proliferation and differentiation^34^. Significantly increased gene expression of PTGS2 has been found in the ectopic and eutopic endometrium of women with endometriosis compared to women without this condition^34,35^. Furthermore, PTGS2 expression in the eutopic endometrium of women with endometriosis has been shown to be significantly higher in those with more severe dysmenorrhea^36^. Targeting key factors involved in endometriosis pathogenesis and symptomatology may contribute to the effectiveness of fenoprofen in treating endometriosis.

Our study has several limitations. The endometriosis signatures were generated from bulk gene data that included 105 microarray samples; they could be made more robust by incorporating additional public expression datasets. Single-cell data could also be used to investigate the potential effects of drug hits on specific types of endometrial cells and for combination therapy predictions. Drugs like ibuprofen and GnRH antagonists that lack representation in the CMAP data used by the drug repurposing pipeline would not be identified by the pipeline, nor would drugs that are represented in CMAP but filtered out during pre-processing due to profile inconsistencies^10^, such as the COX-2 selective inhibitor NSAIDs. The nature of the drug repurposing pipeline prioritizes drugs that have a high reversal effect on the disease signature; it does not take into account whether transcriptional effects are limited solely to genes that the disease also affects. A drug that causes wide-ranging gene changes—including reversal to the gene changes caused by endometriosis—may present in the list of identified drugs, and may be therapeutic. However, unrelated gene changes could cause undesirable side effects, and depending on specificity and severity of side effects, these drugs may have limited applicability clinically. Moreover, in our study, we did not assess the effects of fenoprofen on disease burden. Out of all the drug candidates across every endometriosis signature, many were antipsychotics and other drugs that affect a wide range of genes. In addition, the compounds from the CMap dataset were tested on cancer cell lines; the drug’s effects on endometrial tissue would be far better determiners of its potential applications to endometriosis. A further limitation of our study is that we validated fenoprofen as a potential endometriosis therapeutic in a rodent model. Although our model mimics many disease features of women with endometriosis, rats do not menstruate or develop endometriosis spontaneously. Therefore, menstruating non-human primates could be considered a more appropriate model as they develop endometriosis spontaneously; however, because of their close phylogenetic relationship to humans, these models come with unique ethical considerations^52,53^ as well as limiting financial cost.

To summarize, we applied a computational drug repurposing pipeline to identify potential therapeutics for endometriosis-related pain. The pipeline returned many known treatments as well as novel candidates. We tested the identified therapeutic candidate fenoprofen in an established rat model of endometriosis. We determined that fenoprofen successfully alleviated endometriosis-associated vaginal hyperalgesia, a surrogate marker for endometriosis-related pain. These findings validate fenoprofen as a potential endometriosis therapeutic and suggest the utility of future investigation into additional drug candidates identified.

## Data Availability

Data were obtained from the National Center for Biotechnology Information (NCBI) Gene Expression Omnibus (GEO) Database (series accession number GSE51981)

https://www.ncbi.nlm.nih.gov/geo/query/acc.cgi?acc=GSE51981

## Contributions

D.B, A.B, L.C.G, S.M., and M.S. designed the study, experiments, and analytic plan. T.T.O, A.B., D.B, C.L., and S.M. carried out data acquisition, processing, and analysis. T.T.O, A.B., D.B, B.L.L, I.K., B.G., D.K.S., J.C.I., L.C.G., S.M., and M.S. carried out computational and statistical analysis. T.T.O, A.B., D.B, B.L.L, I.K., B.G., D.K.S., J.C.I., L.C.G., S.M., and M.S. interpreted results. E.A., L.M., L.L., and S.M. carried out the validation experiments and analyzed the relevant data. T.T.O, A.B., and S.M. wrote the manuscript. All the authors participated in relevant discussions, edited and reviewed the manuscript.

## Acknowledgements

The work was in part supported by NIH P01HD106414 (T.T.O., A.B., B.G., D.K.S., J.C.I., L.C.G., S.M., M.S.), NIH P50 HD055764 (A.B., S.H., S.S., W.W., J.C.I., L.C.G., M.S.), and NIH R00HD093858 (E.A., L.M., L.L., S.M.), as well as by the March of Dimes Prematurity Research Center at UCSF (T.T.O., B.L.L., I.K., M.S.), the March of Dimes Prematurity Research Center at Stanford University (B.G., D.K.S.), and the Stanford Maternal and Child Health Research Institute (B.G., D.K.S.). The authors acknowledge the use of resources developed and supported by the UCSF Bakar Computational Health Sciences Institute Information Commons team, and thank members of this team for technical support. The authors also thank the Center for Data-driven Insights and Innovation at UC Health (CDI2; https://www.ucop.edu/uc-health/functions/center-for-data-driven-insights-and-innovationscdi2.html), for its analytical and technical support related to use of the UC Health Data Warehouse and related data assets, including the UC COVID Research Data Set (CORDS).

## Data and Code Availability

Data were obtained from the National Center for Biotechnology Information (NCBI) Gene Expression Omnibus (GEO) Database (series accession number GSE51981) https://www.ncbi.nlm.nih.gov/geo/query/acc.cgi?acc=GSE51981

The UCSF EHR database is available to UCSF-affiliated individuals who can contact UCSF’s Clinical and Translational Science Institute (CTSI) or the UCSF’s Information Commons team for more information. UCDDP is only available to UC researchers who have completed analyses in their respective UC first and have provided justification for scaling their analyses across UC health centers.

Code for transcriptomic data processing associated with the current submission is available at https://doi.org/10.3389/fimmu.2021.788315, and code for computational drug repurposing pipeline associated with the current submission is available at https://doi.org/10.1053%2Fj.gastro.2017.02.039.

