## Supplementary Table 1 for "Applying a computational transcriptomics-based drug repositioning pipeline to identify therapeutic candidates for endometriosis"

| Cycle-Phase | Disease severity | | | Total |
| --- | --- | --- | --- | --- |
| | Control | Stages I-II | Stages III-IV | |
| PE | 20 | 10 | 17 | 47 |
| ESE | 6 | 6 | 12 | 24 |
| MSE | 8 | 8 | 18 | 34 |
| **Total** | **34** | **24** | **47** | **105** |
